# Effectiveness and cost-effectiveness of the Keep-on-Keep-up (KOKU) digital falls prevention programme in community-dwelling older adults: Results of a randomised controlled trial

**DOI:** 10.64898/2026.07.07.26357131

**Authors:** Chloe French, Amelia Parchment, Bolanle Odebiyi, Chunhu Shi, Saima Bashir, Dawn Dowding, Roman Kislov, Alex Thompson, Dawn A Skelton, Margaret Clarke, Yvonne Sylvestre Garcia, Saima Ahmed, Chris Todd, Peter Bower, Emma Stanmore

## Abstract

**Background:** Falls are a leading cause of injury-related hospital admissions among older adults with substantial burden on health and social care systems. Digital exercise programmes may improve physical function at scale and complement traditional services. Keep-On-Keep-Up (KOKU) is an NHS-approved digital programme offering progressive, evidence-based exercises and education on fall prevention. We aimed to evaluate the effectiveness and cost-effectiveness of KOKU for improving balance, physical function and reducing fall risk among community-dwelling older adults.

**Methods:** A two-arm, parallel group randomised controlled trial was conducted with community-dwelling older adults (≥60 years). Participants were randomised (1:1) to receive KOKU alongside standard care (strength and balance exercise advice and a falls prevention leaflet) or standard care alone. The primary outcome was balance function at 12 weeks (Berg Balance Score). Secondary outcomes included lower limb strength, concerns about falling, falls, mood, pain, fatigue, healthcare utilisation, health-related quality of life and usability. A modified intention-to-treat approach was used to analyse effectiveness and cost effectiveness.

**Results:** A total of 202 older adults (mean age 76.8 years, 72.8% female) were enrolled (102 intervention; 100 control). Retention at 12-weeks was 89.1% (91 intervention; 89 control). Compared with standard care, KOKU significantly improved balance function at 12 weeks after adjusting for baseline scores (mean difference: 6.35, 95% CI: 4.48, 8.22). KOKU was associated with lower mean falls related costs (incremental cost: -£62.98, 95% CI -£218.54 to £40.22) and a QALY gain of 0.020 (95% CI 0.003 to 0.035).

**Conclusion:** The KOKU programme improves balance with preliminary evidence of cost-effectiveness among community-dwelling older adults.

**Trial registration number:** NCT06687135

## Introduction

Functional decline, frailty and falls are common concerns associated with ageing and can negatively impact older adults’ independence, mobility, and quality of life [1]. Frailty increases vulnerability to falls and subsequent falls further accelerate functional deterioration [1]. In particular, declining balance, strength and physical function can increase difficulties with activities of daily living and elevate the risk of falls, which are a major cause of physical injury and psychological harm, hospitalisation, and loss of independence in later life. Consequences extend to caregivers and healthcare systems through increased service utilisation, long-term care admissions, and substantial economic costs [2].

Evidence consistently highlights the role of strength and balance training in reducing functional decline [2]. More specifically, regular and progressive exercise programmes that target muscle strength, coordination and postural stability have been shown to improve physical function, support independence and reduce fall rates [3,4]. However, despite the efficacy of such interventions, participation can be low and hindered by logistical, motivational, and environmental barriers such as transportation difficulties or limited programme availability.

The development and implementation of accessible, scalable, and effective interventions that support physical function is an urgent public health priority given the ageing global population and high incidence of falls. Digital health has the potential to augment traditional exercise programmes and overcome some of the barriers to accessibility and personalisation. Preliminary evidence suggests that digital interventions may be both feasible and acceptable for older populations [5], but high-quality research examining their effectiveness in improving balance, function and fall risk remains limited [6].

Keep-on-Keep-up (KOKU) [7,8] is a digitally delivered strength and balance programme. KOKU was co-developed with older adults and falls prevention experts. The development process was informed by MRC guidelines for developing complex interventions [9] and established behaviour change techniques [10,11]. Feasibility trials within and outside of the UK have shown preliminary evidence that KOKU has high usability with improved balance after independent use [12,13]. While these findings are promising, they remain limited by small sample sizes and lack of economic and implementation evaluations.

Therefore, this study aims to evaluate the effectiveness and acceptability of the KOKU programme and provide an estimate of falls related costs and Quality-adjusted life year (QALY) gains. The primary objective was to investigate whether the digital intervention can improve balance compared with standard care among community-dwelling older adults, assessed using the Berg Balance Scale (BBS) [14]. The secondary objectives were to investigate the effectiveness of the KOKU programme on falls risk, physical function, pain, mood, fatigue, healthcare utilisation, health-related quality of life (HRQoL) and fall rate as well as the usability and acceptability of KOKU.

## Methods

### Study design

This mixed-methods two-arm individual randomised controlled trial (RCT) investigated a digital, progressive strength and balance programme (KOKU) plus standard care against standard care alone (strength and balance exercise advice and a falls prevention leaflet). Qualitative data was collected alongside the trial to explore acceptability of the intervention and study processes through a subset of participants through semi-structured interviews and healthcare providers through interviews and focus groups. The trial protocol was published [15] and registered on ClinicalTrials.gov (NCT06687135). Ethical approval was obtained prior to recruitment or data collection, and all participants completed an informed consent form attesting to their voluntary participation in the trial.

### Setting and participants

Participants were community-dwelling older adults aged 60 years and over, able to safely use the tablet-based KOKU programme and mobilise indoors independently with or without a walking aid. Full eligibility criteria are available in Supplementary Table 1.

Potential participants were referred from community settings across the ten localities of Greater Manchester in the Northwest of England. These settings included adult social care teams, housing providers, and charity organisations which aim to support older adults to remain independent in their own homes. Trained research staff checked eligibility of referred participants, explained study procedures, and provided participant information sheets.

### Randomisation and blinding procedure

Randomisation was performed by an independent researcher to allocate participants in a 1:1 ratio to either the intervention or control group, using block randomisation of varying sizes (4, 6, and 8) to prevent researchers from guessing the next participants assigned treatments-thus better minimising the risk of selection bias while maintaining equal group sizes. The random numbers were computer-generated by a statistician not involved in data collection nor in the allocation process. Allocation was not released until after baseline assessments had been completed and the allocation process was secured on Research Electronic Data Capture (REDCap) [16]. Blinding of participants, outcome assessors and those delivering the treatment was not undertaken given the nature of the intervention. Additional details of the randomisation approach and blinding procedures were included in the protocol [15].

### Intervention

Full details of the intervention has been described in the protocol [15] using the TIDieR checklist [17]. In brief, participants allocated to the intervention group received an iPad preloaded with the KOKU digital programme and were instructed to use the programme for 20-30 minutes, three times a week for 12-weeks. Researchers provided initial training at the baseline visit to familiarise participants with the use of KOKU. Participants were asked to follow KOKU instructions for exercises at home and ad hoc technical support was available if required. KOKU offers a progressive strength and balance programme with animated characters (eCoaches) providing instructions, guidance, safety information, automated reminders and in-app progress tracking. Additional details about the types of exercises and development (including use of behaviour change techniques and Patient and Public Involvement (PPI)) have been reported elsewhere [15].

Participants in the intervention group also received home exercise leaflets that were developed based on the Falls Management Exercise (FaME) programme [18,19] and the Otago Exercise Programme (OEP; [20]) (constituting routine or usual care) alongside the KOKU digital programme.

Participants in the control group received the same leaflets without KOKU and were asked to undertake their choice of exercises from those provided in the leaflets three times a week for 12 weeks. After all measurements had been collected, participants in the control group had the opportunity to use the KOKU programme.

### Data collection and analysis

Participant characteristics were recorded at baseline and included age, sex, ethnicity, employment, marital status, fall history, and self-reported vision.

### Outcomes

Researchers completed follow-up assessments of the following outcomes at 6 weeks and/or 12 weeks as previously reported [15]. The primary outcome was balance function at 12 weeks, assessed using the validated BBS [14]. Secondary outcomes included a functional assessment of lower limb strength measured in seconds using the Five Times Sit-to-Stand (5-STS) chair test [21]; HRQoL measured using the European Quality of Life 5 Dimensions (EQ-5D-5L) and visual analogue scale (EQ-VAS) [22]. EQ-5D-5L health states were converted into utility values using the recently published UK value set [23]. Mood was measured using the 5-item Geriatric Depression Scale (GDS; [24]); self-reported physical activity measured using the Physical Activity Scale for the Elderly (PASE; [25]); concerns about falling measured using the Short Falls Efficacy Scale (Short FES-I; [26,27]); and risk of falling measured using the 5-item Falls Risk Assessment Tool ((FRAT) [28]). Visual analogue scores for pain and fatigue were also included ranging from 0 to 100 with higher scores indicating greater pain or fatigue. The primary objective of the economic analysis was to estimate the incremental cost-effectiveness of the KOKU programme compared with usual care. Fall count, severity and resource use data was assessed via self-reported fall calendars for 24 weeks (covering the 12-week intervention and subsequent 12-weeks providing longer-term follow-up), with follow-up phone calls by a researcher (CF). This captured health service utilisation related to falls including ambulance attendances, emergency department visits, GP/doctor consultations, and hospital admissions (public or private) to inform the cost-effectiveness analysis. Unit costs for NHS and Personal Social Services (PSS) resource use were taken primarily from the Unit Costs of Health and Social Care by the PSS Research Unit (2024) [29] and the NHS National Cost Collection [30].

Participants in the intervention group completed questionnaires on the usability and acceptability of KOKU at 12 weeks. Specifically, the System Usability Scale (SUS) [31], the Theoretical Framework for Acceptability (TFA) based on a 5-point Likert scale [32], and the Short Version of the User Experience Questionnaire [33].

### Qualitative interviews and focus groups

After the 12-week intervention, semi-structured interviews or focus groups explored in-depth, qualitative perspectives regarding acceptability and experiences of using the KOKU programme as well as study related processes. A sub-sample of older adults randomised to receive KOKU were recruited purposively with the aim of capturing diversity in gender, geographical area across Greater Manchester, ethnic background and engagement with KOKU. To support reflexive practice and reduce the influence of existing researcher-participant relationships, interviews were conducted by qualitative researchers who had not been involved in delivering KOKU to the included participants. The three researchers involved in interviewing (AP, BO, CF) rotated participants to maintain this separation. Healthcare providers were also recruited purposefully, focusing on those supporting the KOKU trial within their organisation. Sampling aimed to include a broad mix of care provider types and localities across Greater Manchester to reflect the varied organisational contexts of sites involved in the KOKU trial.

Interviews and focus groups were completed online or face-to-face based on the preference of the participant and all were digitally audio recorded and transcribed verbatim.

### Sample size calculation

A sample size calculation (5% two-sided significance level, 90% power) to detect a 5-point improvement in BBS at 12 weeks (based on a standard deviation of 14), including an expected 13% loss to follow up, indicated that 98 participants per group were required [34,35].

### Statistical analyses

Analysis of intervention effectiveness used Stata version 14.0 and cost-effectiveness analysed using Stata version 19.0 [36]. Descriptive statistics for all baseline characteristics were summarised for all randomised participants who started the trial.

Analyses were conducted on a modified intention-to-treat basis, including all randomised participants with baseline and at least one follow-up data point. Continuous outcomes (BBS, EQ-5D-5L, EQ-VAS, PASE, Short FES-I, 5-STS, pain scale, fatigue scale) were analysed using mixed models for repeated measures (MMRM), including fixed effect for group allocation, time point (6 and 12 weeks) adjusting for baseline scores, and interactions of fixed terms with time point. The primary comparison was the contrast (adjusted mean difference) between treatment groups at 12 weeks. Missing data were assumed to be missing at random, which is consistent with the MMRM approach. To retain all valid 5-STS data in the statistical analysis, missing data from participants unable to complete the test were imputed using a fixed value of 61 seconds, consistent with previous studies [37] and based on the upper cut-off in the validated Short Physical Performance Battery (SPPB) [38].

Binary logistic regression models with robust standard errors were fitted separately for each outcome and timepoint (6 and 12 weeks) to investigate the proportion of participants at risk of depression (GDS ≥ 2), having high concerns about falling (short FES-I ≥ 11), or being at a high fall risk (FRAT ≥ 3) based on established thresholds [24,26,28,39]. All models were adjusted for baseline values of the respective outcome. Fall rates were compared between groups using Poisson regression models, an established method for analysing count data and event rates [40]. Results are presented as incidence rate ratios (IRRs) with 95% confidence intervals adjusting for baseline values.

Usability and acceptability data from participants receiving the intervention was summarised descriptively. Data from the SUS was compared with the industry average of 68 and the recommended cut-point that products scoring >80 are considered to have excellent usability [41]. Data from the user experience questionnaire was compared with a benchmark dataset containing 21175 individuals from 468 studies [42].

### Economic analysis

For the cost utility analysis, the outcome was QALYs accrued from baseline to the 6- and 12-week follow-up (calculation detailed in Supplementary Material). Health states were measured using the EQ-5D-5L [23,43].

The cost of delivering the intervention was estimated by identifying key resources: KOKU training for care providers, licensing fee, vendor implementation cost, post-implementation support, and digital tablets (e.g. iPads or Android Tablets) to run the digital programme (detailed in Supplementary Material). As the economic evaluation adopts a health and social care perspective, patient-borne and indirect societal costs (e.g. lost productivity) were excluded [44].

Cost-effectiveness of the KOKU programme was evaluated over the 12-week trial horizon, from the NHS and PSS perspectives. As the time horizon was 12 weeks, discounting of costs and outcomes was not applied.

Differences in costs and QALYs between KOKU and control groups were estimated using regression analysis. Incremental costs were modelled using a generalised linear model (GLM) with Gaussian family and identity link, whereas incremental effects were estimated with a β-distributed GLM with a logit-link to account for the bounded distribution of QALYs. Models included intervention indicator, baseline cost, EQ-5D-5L utility, age, ethnicity, and study stratification indicators as fixed effects.

Parameter uncertainty was explored using non-parametric bootstrapping with 10,005 replications across the imputed datasets (667 bootstrap replications per imputed dataset). For each replication, incremental costs, incremental QALYs, incremental cost-effectiveness ratios (ICERs), and incremental net monetary benefits (INMBs) were estimated at willingness-to-pay thresholds of £20,000 and £30,000 per QALY gained [45].

Joint uncertainty in costs and outcomes was illustrated using cost-effectiveness planes. Cost-effectiveness acceptability curves (CEACs) were generated to estimate the probability that the KOKU programme was cost-effective across alternative willingness-to-pay thresholds. In addition, the distribution of incremental net monetary benefit was presented graphically to further characterise decision uncertainty.

Sensitivity analyses were conducted to assess the robustness of the results to alternative assumptions: (1) complete case analysis; (2) excluding tablet costs by assuming that all participants had access to their own tablet; (3) varying published unit costs for health and social care by ±10%; and (4) assuming that one in four participants required a tablet, corresponding to an intervention cost of £20 per patient.

### Qualitative analysis

Following Braun and Clarke’s six-phase approach [46,47], Reflexive thematic analysis was employed both inductively and deductively to explore patterns of meaning across participant and staff interviews. A qualitative researcher (AP), who was also involved in the delivery of KOKU and the interviewing of participants (1) familiarised herself with the data through immersion in the interview transcripts; (2) generated initial codes to reflect meaningful data relevant to the research questions. Both semantic and latent coding was used to explore explicit, surface meaning communicated by participants in addition to deeper layers of meaning and underlying assumptions; (3) developed initial themes to capture patterns of shared meaning across the interview dataset; (4) reviewed and refined themes iteratively.

Discussion with other members of the research team as the analytical process progressed supported reflexive practice and encouraged critical reflection on the assumptions and positionality shaping the qualitative findings. This was important given that the lead qualitative researcher was an adult without lived experience of mobility issues or chronic health conditions, which may influence interpretation of data and construction of findings.

## Results

Between July 2024 and March 2025, 372 older adults were assessed for eligibility, 205 were randomised, including three who were randomised in error as they did not meet eligibility criteria. 202 older adults were enrolled onto the trial (102 intervention, 100 control) (Figure 1) [48].

**Figure 1:**
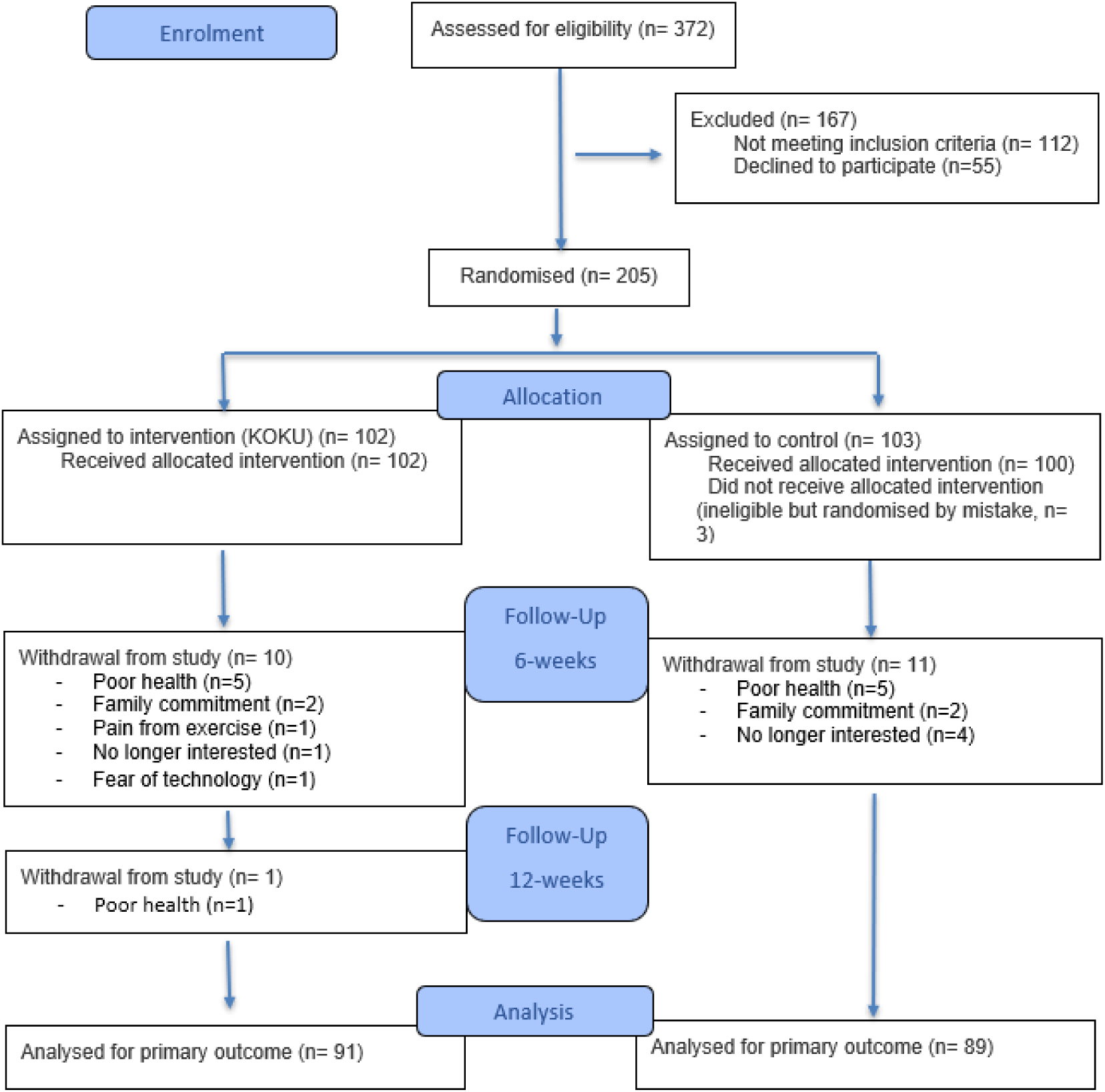
Flowchart of participants throughout the study.

Retention of participants was 89.1% at 12-weeks, most participants who withdrew from the study did so in the first 6-weeks of enrolment. Withdrawal was balanced across study arms (Figure 1). Reported reasons for withdrawal in both groups were primarily poor health (n= 10) or family commitments (n= 4), with no evidence that dropout was related to treatment allocation or study outcomes. In addition, baseline characteristics (socio-demographics and health status) were comparable between those who withdrew and those who completed follow-up, with slightly better retention in ethnic minority groups and those who were single or never married (Supplementary Table 2).

Baseline characteristics of participants are displayed in Table 1. The mean age was 76.8 years (range 60-98 years); a large proportion were female (72.8%) and of White British ethnicity (85.6%). Almost all participants (93.6%) took at least one medication and 72.8% took ≥4 medications. Slightly more participants randomised to receive KOKU had self-reported no falls in the previous 12 months (61.8% intervention, 55.0% controls) (Table 1).

**Table 1:** Demographic and baseline characteristics of participants by intervention group.

|  | **Intervention**<br>**(n= 102)** | **Control**<br>**(n= 100)** | **Total**<br>**(n= 202)** |
| --- | --- | --- | --- |
| **Age, mean (SD)** | 76.4 (8.1) | 77.1 (8.9) | 76.8 (8.5) |
| **Sex, n (%)** |  |  |  |
| Male | 24 (23.5) | 31 (31.0) | 55 (27.2) |
| **Ethnicity, n (%)** |  |  |  |
| White British | 86 (84.3) | 87 (87.0) | 173 (85.6) |
| Black or Black British | 11 (10.8) | 5 (5.0) | 16 (7.9) |
| Other | 5 (4.9) | 8 (8.0) | 13 (6.5) |
| **Place born, n (%)** |  |  |  |
| UK | 86 (84.3) | 82 (82.0) | 168 (83.2) |
| Other | 16 (15.7) | 18 (18.0) | 34 (16.8) |
| **Marital status, n (%)** |  |  |  |
| Single, never married | 18 (17.6) | 8 (8.0) | 26 (12.9) |
| Married/ living with a partner | 37 (36.3) | 34 (34.0) | 71 (35.1) |
| Divorced | 15 (14.7) | 11 (11.0) | 26 (12.9) |
| Separated | 1 (1.0) | 2 (2.0) | 3 (1.5) |
| Widowed | 31 (30.4) | 45 (45.0) | 76 (37.6) |
| **Device ownership/ access\*, n** |  |  |  |
| Tablet | 68 | 54 | 122 |
| Smartphone | 91 | 82 | 173 |
| Laptop | 45 | 42 | 87 |
| **Takes medication, n (%)** | 94 (92.2) | 95 (95.0) | 189 (93.6) |
| **4 or more medications n (%)** | 74 (72.5) | 73 (73.0) | 147 (72.8) |
| **Fractured a bone, n (%)** |  |  |  |
| Yes | 48 (47.1) | 51 (51.0) | 99 (49.0) |
| No | 54 (52.9) | 49 (49.0) | 103 (51.0) |
| **Recently had surgery, n (%)** |  |  |  |
| Yes | 8 (7.8) | 10 (10.0) | 18 (8.9) |
| No | 94 (92.2) | 90 (90.0) | 184 (91.1) |
| **Self-reported eyesight, n (%)** |  |  |  |
| Excellent | 11 (10.8) | 10 (10.0) | 21 (10.4) |
| Good | 69 (67.6) | 55 (55.0) | 124 (61.4) |
| Fair | 19 (18.6) | 27 (27.0) | 46 (22.8) |
| Poor | 3 (2.9) | 5 (5.0) | 8 (4.0) |
| Very poor/ Partially sighted/<br>Blind | 0 (0) | 3 (3.0) | 3 (1.5) |
| **Fall (last 12 months), n (%)** |  |  |  |
| Never | 63 (61.8) | 55 (55.0) | 118 (58.4) |
| Once | 17 (16.7) | 21 (21.0) | 38 (18.8) |
| Twice or more | 22 (21.6) | 24 (24.0) | 46 (22.8) |
| **EQ-5D, mean (SD)** | 0.77 (0.25) | 0.69 (0.29) | 0.73 (0.27) |
| **EQ-VAS, mean (SD)** | 70.26 (18.12) | 64.87 (19.53) | 67.59 (18.98) |
| **Risk of depression, n (%)** |  |  |  |
| Low risk (GDS<2) | 60 (58.8) | 61 (61.0) | 121 (59.9) |
| At risk (5-GDS≥2) | 42 (41.2) | 39 (39.0) | 81 (40.1) |
| **PASE, mean (SD)** | 71.67 (44.47) | 70.38 (45.01) | 71.02 (44.63) |
| **Short FES-I, mean (SD)** | 14.09 (5.80) | 15.64 (6.14) | 14.86 (6.01) |
| **FRAT, n (%)** |  |  |  |
| Lower risk of falling (0-2) | 63 (61.8) | 52 (52.0) | 115 (56.9) |
| Greater risk of falling (≥3) | 39 (38.2) | 48 (48.0) | 87 (43.1) |
| **BBS, mean (SD)** | 42.97 (10.51) | 42.39 (9.95) | 42.68 (10.21) |

*\*Frequencies are greater than sample size given that some participants owned multiple types of technology. EQ-5D-5L- EuroQol 5-Dimension 5-level; EQ-VAS- EuroQol visual analogue scale; GDS- Geriatric depression score; Short FES-I- International short falls efficacy scale; FRAT- Falls risk assessment tool; BBS- Berg balance scale*

### Primary outcome

At 12-weeks, participants receiving KOKU (n=91) had a higher mean BBS score (indicating greater balance function) than participants receiving standard care (n=89), with a between group difference of 6.35 points (95% CI: 4.48, 8.22; Table 2). There was strong evidence that the KOKU intervention improved BBS scores at both 6 and 12 weeks. The treatment-by-time interaction was statistically significant (p<0.001), indicating that the effect of the intervention increased over time, with a larger improvement observed at 12 weeks than at 6 weeks.

**Table 2:**
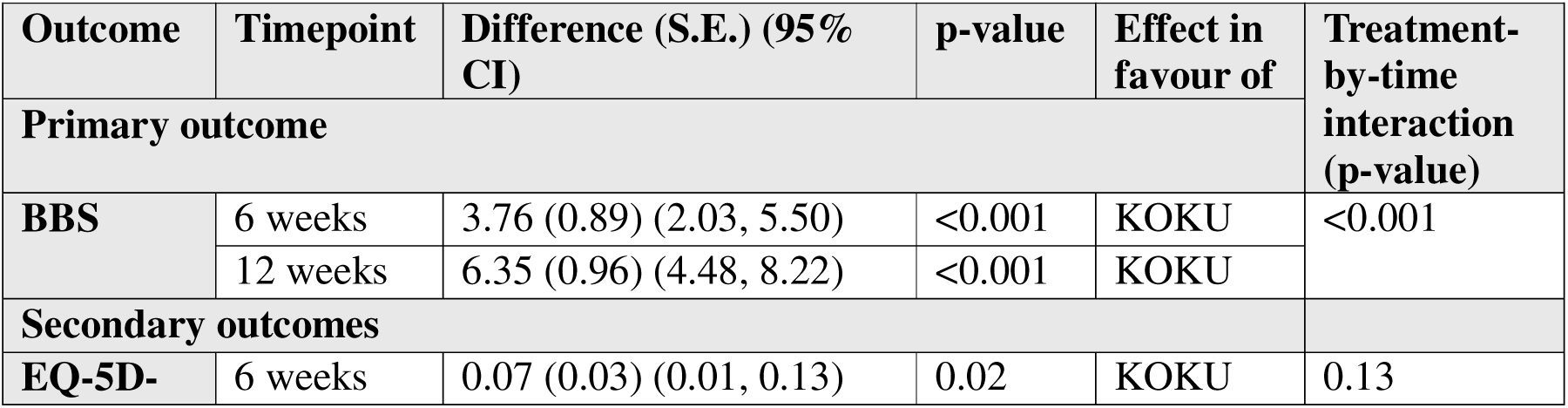
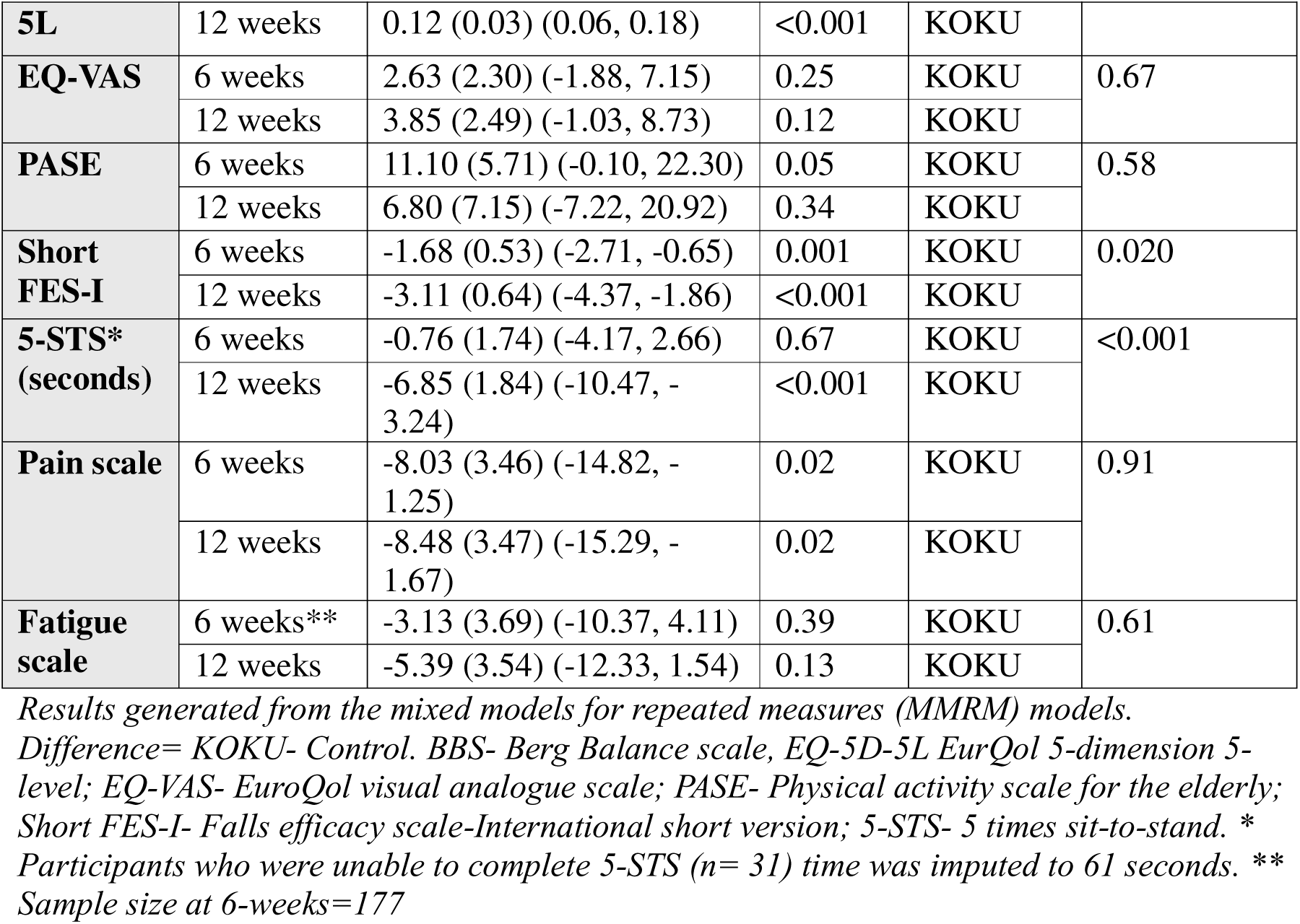
Treatment effects on continuous outcomes at 6-weeks (n=178) and 12-weeks (n=180).

### Secondary outcomes

The mean and SD of outcomes reported in Table 2 are available in Supplementary Table 3 split by treatment group at 6- and 12-weeks. There was a significant difference in health-related quality of life assessed by the EQ-5D-5L at both 6- and 12-weeks between participants who received KOKU and those who received usual care adjusting for baseline values (Table 2). Participants receiving the KOKU intervention reported better quality of life based on fewer problems with mobility, self-care, usual activities, pain and anxiety (Supplementary Table 4).

There was no difference in overall health perception assessed using EQ-VAS or physical activity level from PASE at 6 or 12 weeks. Participants who received the KOKU intervention had less concern about falling (assessed using short FES-I) at both 6- and 12-weeks compared to participants in the control group (mean difference: −3.11 at 12 weeks; 95% CI: −4.37, 2.66; Table 2). In addition, participants receiving the KOKU intervention had improved (lower) 5-STS time (seconds) at 12-weeks compared to participants in the control group (mean difference: −6.85; 95% CI: −10.47, −3.24).

At 12-weeks, participants who received the KOKU intervention had lower concerns about falling (short FES-I score <11) compared to participants in the control group after adjusting for baseline short FES-I scores (aOR=0.34, 95% CI: 0.17, 0.70, p<0.01, Table 3). Table 3 shows that the KOKU intervention did not have a statistically significant effect on reducing risk of falls (FRAT<3) or depression (GDS<2) at 6 or 12-weeks when accounting for their respective baseline values. A total of 73 falls (52% control; 48% KOKU) were self-reported during the 24-week follow-up (IRR: 0.84, 95% CI: −0.63, 0.28) showing no significant difference in number of falls between groups.

**Table 3:** Binary logistic regression showing adjusted odds ratio at 6- and 12-weeks comparing intervention (KOKU) and control groups.

|  | Number of participants |  | aOR (95% CI) | p-value |
| --- | --- | --- | --- | --- |
|  | KOKU | Control |  |  |
| High concern of falling (Short FES-I <11) |  |  |  |  |
| 6-weeks | 91 | 87 | 0.74 (0.36, 1.52) | 0.42 |
| 12-weeks | 91 | 89 | 0.34 (0.17, 0.70) | <0.01 |
| High risk of falling (FRAT <3) |  |  |  |  |
| 6-weeks | 91 | 87 | 0.63 (0.32, 1.27) | 0.20 |
| 12-weeks | 91 | 89 | 0.58 (0.26, 1.27) | 0.17 |
| High risk of depression (GDS<2) |  |  |  |  |
| 6-weeks | 91 | 87 | 0.59 (0.27, 1.28) | 0.18 |
| 12-weeks | 91 | 89 | 0.65 (0.30, 1.40) | 0.27 |

*aOR- adjusted odds ratio; Ref- Reference value; Short FES-I: Falls Efficacy Scale International (short version); FRAT- Falls Risk Assessment Tool; GDS- Geriatric Depression Scale*

Results from the SUS show that KOKU had excellent usability (median score 88.75, IQR: 77.50, 100.00, n= 90). The seven constructs of the theoretical framework for acceptability (TFA) were also considered, and general acceptability was high (mean score 4.64; SD: 0.48). Supplementary Table 5 presents the mean score for each of the seven constructs. In addition, participants using KOKU (n=90) reported excellent user experience across pragmatic quality, hedonic quality and overall (Supplementary Figure 1) [42].

### Health economic outcomes

QALYs were missing for 25/202 (12.4%) participants. Missing outcomes were imputed using multiple imputation (15 imputations). We applied chained equations (MICE) with predictive mean matching, combined using Rubin’s rules [49,50]. The imputation model included the intervention indicator, baseline utility values, total cost and key demographic covariates.

The estimated cost to deliver the 12-week KOKU programme was £43 per person (Supplementary information). Overall, the largest contributors to total cost were the licensing fee (£24) and digital tablets (£16) required to run the programme (based on one in five participants in intervention arm requiring a tablet).

Unadjusted EQ-5D-5L utilities for both groups across the follow up are shown in Figure 2, indicating utility gains in both arms, with a larger improvement in the intervention group. The unadjusted between-group difference in mean utility was 0.178 [12-week QALYs difference is 0.031]. Supplementary Table 6 reports primary and secondary healthcare unit costs.

**Figure 2:**
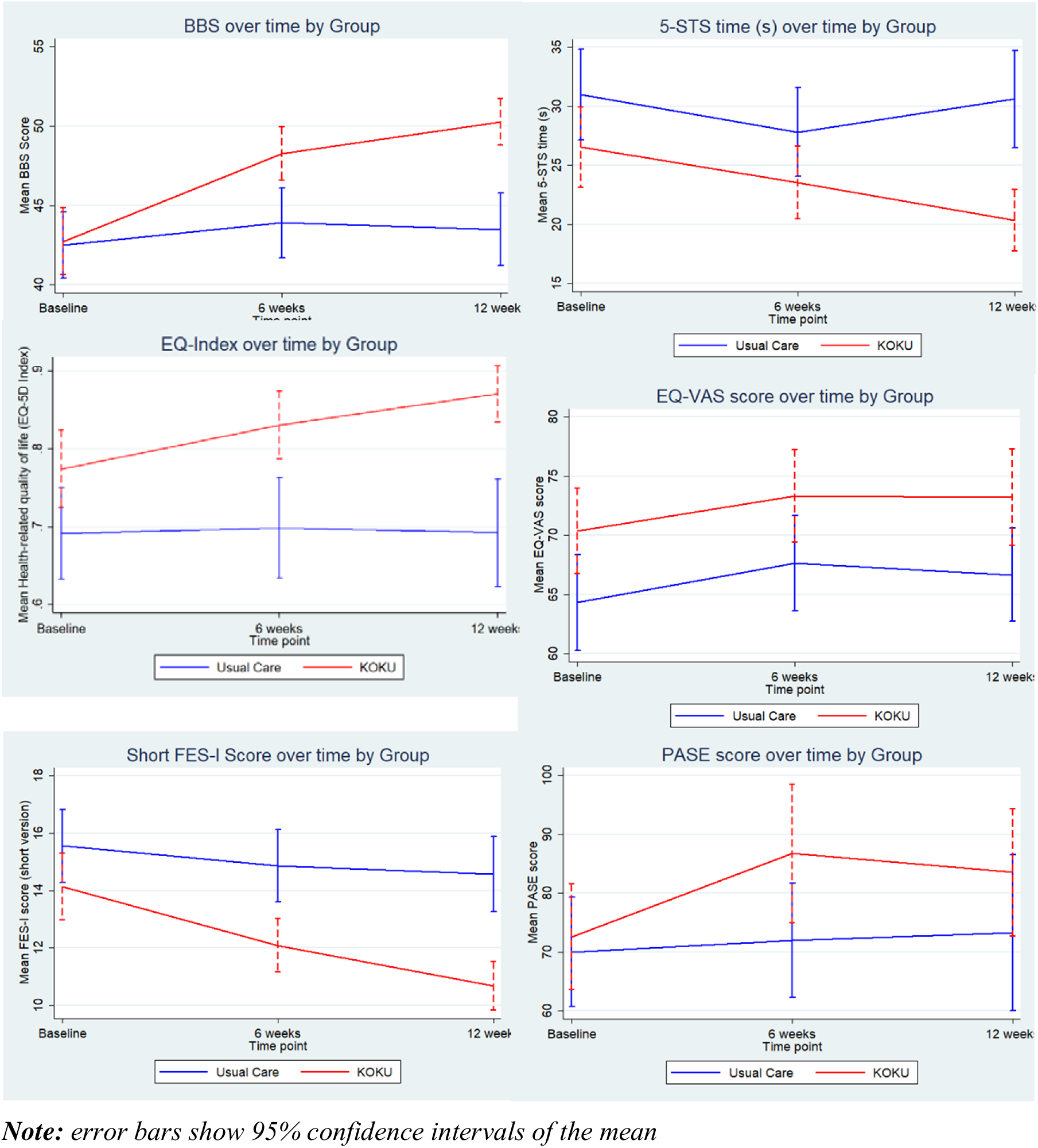
Outcome measures between participants receiving KOKU and usual care over time.

The incremental analysis results are presented in Table 4. The KOKU programme was associated with an adjusted incremental fall-related cost of -£62.98 (95% CI -£218.54 to £40.22) and an average incremental QALY gain of 0.020 (95% CI 0.003 to 0.035) over the 12-week period. QALY gain was statistically significant, while the cost difference was not statistically significant, although the point estimate favoured cost savings. As the intervention was both less costly and more effective than usual care, it was considered dominant and therefore no ICER was calculated.

**Table 4:** Cost-effectiveness analysis comparing KOKU with control group.

|  | Coeff. | Bootstrapped SE | Bootstrapped 95% CI |
| --- | --- | --- | --- |
| Incremental cost (£) | -62.98 | 68.18 | [-218.54, 40.22] |
| Incremental QALYs | 0.020 | 0.008 | [0.003, 0.035] |
| ICER (£) | Dominant |  |  |

**Note:** SE = standard error; CI = confidence interval; QALY = quality-adjusted life year; ICER = incremental cost-effectiveness ratio. 95% confidence intervals for incremental costs and QALYs were estimated using non-parametric bootstrapping with 10,005 replications (667 replications per multiple imputation dataset). The ICER is reported as dominant where the intervention is both less costly and more effective than the control group).

Cost-effectiveness plane (Supplementary Figure 2) displays the 10,005 bootstrap replications of incremental cost and QALYs, illustrating probabilistic uncertainty around the point estimates. The distribution places a clear majority of draws in the south-east (SE) quadrant, indicating a high probability that the intervention is less costly and more effective. A smaller proportion in the north-east (NE) quadrant shows that, when costs are higher, KOKU generally still yields QALY gains. We estimated Incremental Net Monetary Benefit (INMB) using bootstrap simulations at willingness-to-pay thresholds of £20,000 and £30,000 per QALY. The distributions are entirely above zero, indicating the intervention is likely cost-effective, with higher INMB (greater benefit) at the £30,000 threshold (Supplementary Figure 3).

The cost-effectiveness acceptability curve (Supplementary Figure 4) demonstrates the probability that the KOKU programme is cost-effective rising with the willingness to pay (WTP). The programme is cost-effective even at very low thresholds, well below the NHS lower bound (£20,000). KOKU was likely to be cost-effective in over 90% cases if decision makers are willing to pay approximately £2000 for one additional QALY.

### Sensitivity Analysis

Across sensitivity analyses, incremental fall-related costs remained lower (-£52.38 to -£87.33) and QALYs remained higher (0.019-0.021), compared with usual care, with the intervention remaining dominant under all assumptions (Supplementary Table 7), demonstrating that the cost-effectiveness results were robust to alternative assumptions regarding missing data, unit costs, and tablet provision.

### Qualitative views and perspectives

Thirteen older adults and ten social care staff engaged in qualitative interviews and focus groups to explore their acceptability of KOKU, as well as their perceptions and experiences using it, in order to help conceptualise the intervention’s effectiveness. Twenty interviews and one focus group (with three social care staff from the same organization) were conducted. Characteristics of these participants are displayed in Supplementary Tables 8a, and 8b. Key themes developed during reflexive thematic analysis are presented below with relevant quotes provided in Supplementary Table 9.

### Older adults’ perspectives

#### Theme 1: High ease of use increases confidence, motivation and engagement

Most participants found KOKU easy to use and highlighted that the programme appeared to have been designed with and for older adults, including those who were not familiar or experienced with digital devices and those with different health needs.

The structure of the programme was considered simple to follow and beneficial for older adults, including those with lower levels of digital literacy and mobility, enabling them to progress at their own pace through the exercises. Participants also highlighted the guided support throughout the programme. The demonstration of exercises by the eCoach, for example, enhanced KOKU’s usability and increased both the confidence and motivation of some participants to engage in and complete prescribed exercises. Some reflected on how the exercises had become easier, more habitual and embedded within day-to-day life over time, leading to long-term maintenance. High ease of use and flexibility of KOKU’s delivery meant that participants were able to engage with the programme at their own convenience, leading to some completing exercise sessions daily or even twice per day.

Some participants did, however, face technical glitches when progressing through KOKU exercises, reducing usability and consequently impacting motivation to continue with the programme. Others experienced confusion regarding the dose of each exercise and felt that counters, timers or guidance on the number of required repetitions could improve usability of the programme, confidence in exercise progression and overall engagement.

#### Theme 2: Improving health, wellbeing and functioning through routine KOKU use

All participants emphasised physical and psychological benefits through regularly engaging with the KOKU programme and in the study itself. Improvements were observed in strength, balance, mobility, energy levels, mood, pain, sedentary behaviour, concerns about falling, confidence in walking and engaging with other activities. One participant discussed an observed reduction in falls since completing the programme. Notably, taking part in the study and using KOKU routinely gave some individuals a sense of purpose. Other participants suggested it should be readily available and offered to *“anybody having any problems with their stability, mobility and low mood” (Participant 12)* and those with specific chronic health conditions that affect balance. Participants similarly reflected on KOKU’S health literacy games as being simple but effective, raising awareness of factors linked to health, wellbeing and independence in later life, with some reporting improvements in diet, hydration and falls awareness. Most participants reported that they would highly recommend KOKU to others, given the benefits they had observed for themselves.

The end of the intervention period was associated with a decline in physical functioning, underscoring the importance of maintaining KOKU to support longer term outcomes.

#### Theme 3: Tailoring challenge to ability

Finally, despite emphasising that KOKU was suitable for older adults, several participants reflected on its level of challenge for those that were more active, independent and engaging in different forms of activity. Providing the option of more difficult exercises for higher functioning users was recommended to ensure all participants were appropriately challenged and able to continue making progress throughout the intervention. Similarly, further progression and the addition of more challenging health literacy games were highlighted as an opportunity for meeting the needs of older adults with higher or increasing levels of knowledge relating to health, wellbeing and functioning.

### Care providers’ perspectives

Three key themes were developed during analysis of interviews with staff supporting KOKU use and study procedures.

#### Theme 1: Simple, safe and beneficial

Similar to the older adult participants, care providers reflected on the high ease of use of KOKU. They perceived KOKU to be highly accessible for both the older adults and their carers, who also benefited from an increased awareness of strength and balance exercises and health literacy around fall prevention.

The programme was considered easy to navigate and follow, with the use of bright contrasting colours, large icons and clear instructions within the programme, supporting usability. They reported that KOKU appeared to have clearly been co-designed with older adults and the embedded, regular safety instructions were noted as an important function. Older adults with some cognitive impairment or extra support needs were perceived as being still able to benefit from KOKU with additional guidance or assistance.

#### Theme 2: Making falls prevention accessible

Staff suggested KOKU aligned strongly with national falls prevention agendas in addition to local and national digital inclusion priorities. One member of staff commented on how engaging in the KOKU study (with iPads/Tablets provided) had enabled older adults facing high socioeconomic deprivation to learn digital skills and take part in an intervention that could benefit their health and wellbeing that may not have otherwise been accessible to them. The use of a digital programme to support falls prevention was additionally highlighted as being highly valuable for older adults that would not find community exercise groups desirable or for those who are homebound. The interactivity of the programme, gamification and embedded behaviour change techniques were felt to increase engagement and motivation of the older adults in completing their daily strength and balance exercises and health literacy games.

Staff commented on the high credibility of KOKU, due to its use of evidence-based content and support from clinicians and some commissioners. This credibility led leadership staff to support the study and endorse the programme amongst other staff and carers. Some discussed increasing digital accessibility by increasing compatibility of KOKU across a range of digital devices including mobile phones and TVs. This recommendation was highlighted as a way to enhance reach and support digital inclusion so that more older adults could benefit from KOKU.

Staff also provided suggestions on how the programme could be adapted and improved for a more diverse range of older adults. Several members of staff felt that KOKU currently only targeted White Caucasian users. Cultural adaptation was one suggested future direction to enhance inclusivity, acceptability and accessibility for ethnically diverse and non-English speaking older adults.

#### Theme 3: Empowering self-management to reduce NHS demand

The overriding value proposition noted by care providers was the use of KOKU for older adults with a range of chronic health conditions and their carers to improve strength, balance and health literacy around falls risk. KOKU was considered to be a digital tool that had potential for reducing pressure on NHS services.

Staff highlighted how, through KOKU use, older adults appeared to self-manage symptoms such as pain, fatigue and low mood better. Staff reported that, for some older adults, simply moving more led to improvements in physical health and psychological wellbeing and gave them increased confidence to take control of their own health. Through taking part in the KOKU study, some care providers reported that service users gained more confidence to join community-led activity groups, supporting overall wellbeing as well as social connectedness. This was particularly important for those who struggled with social isolation and loneliness. Staff members also highlighted that group KOKU sessions could increase confidence, motivation and adherence for some older adults with different cognitive and physical needs, as well as providing important social benefits for those experiencing isolation and loneliness.

## Discussion

This 12-week RCT demonstrates that KOKU, a home-based and progressive digital programme, was effective in improving balance (BBS), lower-limb physical function (5-STS) [51], and concerns about falling (Short FES-I) [26], with observed changes meeting or exceeding established minimal clinically important difference thresholds, alongside improvements in health-related quality of life (EQ-5D-5L) among older adults. These outcomes are widely recognised as key indicators of functional ability and closely linked to fall risk, independence and healthcare utilisation. There was no change, over the 12-week period, in mood, physical activity, fatigue or fall rate. However, the low cost of delivering KOKU (£43 per participant) and preliminary evidence of QALY gains (0.027 difference) suggest that the programme is cost-effective for improving quality of life for health and social care services.

Interviews with older adults combined with questionnaires assessing usability and acceptability highlight that KOKU was simple to use, acceptable as a digital intervention and provided a sense of purpose. Health care providers reported the benefits of providing an alternative or complementary option to face-to-face exercise classes for those unwilling or unable to attend group classes, to provide people who may be within long waiting lists, and the potential to reduce pressures within a healthcare setting.

Although challenges associated with digital exclusion remain, there is a growing acceptability around digital health amongst this population, accelerated by the Covid-19 pandemic [52]. Moreover, policy trends are promoting a shift from analogue to digital services and from treatment to prevention [53]. KOKU was developed with and for older adults; this person-based approach reduces barriers to access and may improve confidence to use digital tools in line with national digital inclusion priorities [54]. Devices and technical support were provided in this study but the potential for digital exclusion is an important consideration for wider implementation. Without appropriate consideration, digital interventions have the potential to risk compounding existing health inequalities by preferentially benefitting those who are already confident and able to engage. Future implementation involves engaging stakeholders including caregivers, community organizations and healthcare professionals to provide ongoing digital support and support sustained engagement. The KOKU programme may help to reduce pressure on face-to-face services and waiting lists by providing an accessible and scalable alternative. In addition, digitally delivered interventions may improve reach among individuals in rural or underserved areas where access to strength and balance classes or falls specialist services are limited.

### Strengths and limitations

Strengths of this study include a robust RCT design, high retention (particularly within ethnic minority groups), and comprehensive measurement of both functional and patient-reported outcomes. In addition, the KOKU platform has been co-designed with older adults, is based on an evidence-based progressive exercise programme, and iPads along with digital support were provided to facilitate digital inclusion. Furthermore, implementation strategies with in-depth qualitative findings are ongoing to support continued use of KOKU among diverse ethnic groups through collaboration with community providers supporting underserved groups. Retention at 12 weeks was high (89.1%) despite recruiting older adults with high baseline medication use (72.8% taking ≥4 medications) and prior fracture history (49%). This indicates that sustained participation was achievable, supporting the feasibility and acceptability of digital interventions among this population.

However, several limitations warrant consideration. The sample was predominantly composed of white, older women so it would be important to examine how we could encourage more men and older people from ethnic minorities to join research studies. Lack of blinding of participants and assessors may have influenced self-report outcomes. The study did not collect usage data (i.e number of sessions completed, or time spent engaged with the KOKU programme) and so changes over time cannot be linked to adherence or engagement with KOKU. Dropout rates were low and balanced between groups, a modified intention-to-treat analysis was employed, and clinically meaningful effect sizes with statistically significant outcomes were maintained, reducing the likelihood that drop-out substantially affected the study findings. Furthermore, resource use and cost data were collected only in relation to falls-related healthcare contacts; broader healthcare utilisation unrelated to falls was not captured, which may underestimate the full economic burden and limit the generalisability of the cost findings. Future work should aim to capture adherence data through a data dashboard or google analytics. Going forward it would also be beneficial to examine whether sustained KOKU engagement (>12 weeks) translates into reduced falls and healthcare utilisation. Future research should involve a longer-term study that is powered to evaluate the effectiveness of KOKU in reducing fall incidence, particularly given that National Institute for Health and Care Excellence (NICE) falls prevention guidelines recommend programmes of at least 12 weeks to achieve sustained benefit [55].

### Conclusion

KOKU demonstrates that relatively low-cost, scalable digital solutions can improve outcomes that are meaningful to older adults, including balance, lower-limb strength, concerns about falling, and perceived quality of life. These findings support the rationale to integrate digital fall prevention strategies into routine care, with the potential to mitigate some of the physical and psychosocial consequences associated with functional decline and falls risk among ageing populations.

## Supporting information

CONSORT

## Data Availability

Supplementary data mentioned in the text are available upon request

## Notes

### Competing Interest Statement

CT is funded by the National Institute for Health and Care Research, Policy Research Unit in Healthy Ageing (reference NIHR206119). PB, DD, JD, RK, ES and CT are co-investigators on the National Institute for Health and Care Research Applied Research Collaboration-Greater Manchester (NIHR200174). AP, SB, CS, SA are funded by NIHR Applied Research Collaboration-Greater Manchester (NIHR200174). The views expressed are those of the authors and not necessarily those of the NHS, the NIHR, the Department of Health and Social Care, or its partner organisations. DS is Director of Later Life Training Ltd, a not-for-profit organisation that delivers training for face-to-face falls prevention delivery, and ES is Director of KOKU Health Ltd.

### Clinical Trial

NCT06687135

### Clinical Protocols

https://www.researchprotocols.org/2026/1/e78840

### Author Declarations

Approvals from The University of Manchester Research Ethics Committees (Ref: 2024-18620-35607) were obtained before commencing participant recruitment.

### Funding

- NHS England: Ref: 31088
- National Institute for Health and Care Research, Applied Research Collaboration- Greater Manchester: NIHR200174

