## Supplementary information for "Effectiveness and cost-effectiveness of the Keep-on-Keep-up (KOKU) digital falls prevention programme in community-dwelling older adults: Results of a randomised controlled trial"

**Supplementary File**

**Supplementary information 1**

QALYs were calculated using time-weighted linear interpolation of utility scores at baseline, 6 weeks, and 12 weeks as follows:

$$QALY=\left[ \frac{U_{baseline}+U_{6-weeks}}{2} \right]\times\frac{6}{52}+ \left[ \frac{U_{6-weeks}+U_{12-weeks}}{2} \right]\times\frac{6}{52}$$

where U denotes the EQ-5D-5L utility score.

**Supplementary information 2**

The estimated cost of delivering the KOKU programme was calculated by identifying all intervention components and apportioning shared costs across users to derive a per-participant intervention cost. Training costs were based on 1 hour of training per carer, with one carer supporting approximately five participants (20 carers for approximately 100–102 participants), valued using the national median hourly wage for care workers (£10.65/hour; Skills for Care, 2022–23), resulting in a cost of approximately £0.18 per participant. Vendor implementation support was estimated from four 15-minute support calls per carer (60 minutes total), costed using the same wage rate and apportioned across participants, contributing a further £0.18 per participant. Licensing costs were estimated at £24.00 per participant. Technical support costs were derived from a £200 service contract with a digital health technology provider and, assuming that one in five users required support, contributed approximately £2.00 per participant. Device costs were included to account for participants requiring access to a tablet to use the application. Assuming that one in five participants required a tablet and based on an amortised tablet cost of £319, the estimated device cost was £16.27 per participant. The total intervention delivery cost was therefore estimated at £42.63 per participant, which was rounded to £43 per participant for the economic analysis.

**Supplementary table 1:** Eligibility criteria of the Keep-on-Keep-up RCT

| **Inclusion criteria** | **Exclusion criteria** |
| --- | --- |
| Willing and able to give informed consent | Those who are currently using digital technologies to exercise |
| Speak English | Currently taking part in an exercise programme or undertaking physiotherapy |
| Able to see and safely use the tablet-based KOKU programme | Those who are unable to comprehend the study procedures |
| Can read instructions with or without glasses, as assessed by the trained research staff | Those who have medical contraindications to exercise due to unstable health e.g. acute illness, severe congestive cardiac failure, uncontrolled hypertension, recent fracture or surgery; Myocardial Infarction or Stroke in past six months; severe cognitive impairment; orthopaedic surgery in last six months, or on waiting list to have orthopaedic surgery; wheelchair users; severe auditory or visual impairment; or other uncontrolled medical conditions likely to compromise the ability to exercise. |
| Are able to mobilise indoors without the help of another person, with or without a walking aid (self-reported) |  |

**Supplementary Table 2**: Comparison of baseline characteristics between total sample and participants who withdrew

|  | **Total sample (n=202)** | **Withdrawals (n=22)** |
| --- | --- | --- |
| Age, median (IQR) | 77.7 (69.6, 82.9) | 66.2 (64.2, 82.0) |
| Sex, n (%)  Male | 55 (27.2) | 6 (27.3) |
| Ethnicity, n (%)  White British  Black or Black British  Other | 173 (85.6)  16 (7.9)  13 (6.5) | 21 (95.5)  1 (4.5) |
| Place born, n (%)  UK  Other | 168 (83.2)  34 (16.8) | 19 (86.4)  3 (13.5) |
| Marital status, n (%)  Single, never married  Married/ living with a partner  Divorced  Separated  Widowed | 26 (12.9)  71 (35.1)  26 (12.9)   3 (1.5) 76 (37.6) | 1 (4.5)  10 (45.5)  5 (22.7)  1 (4.5)  5 (22.7) |
| Device ownership/ access* n  Tablet  Smartphone  Laptop | 122  173  87 | 12  19  12 |
| Takes medication, n (%) | 189 (93.6) | 20 (90.9) |
| 4 or more medication, n (%) | 147 (72.8) | 17 (77.3) |
| Fractured a bone, n (%)  Yes  No | 99 (49.0)  103 (51.0) | 15 (68.2)  7 (31.8) |
| Recently had surgery, n (%)  Yes  No | 18 (8.9)  184 (91.1) | 0  22 (100.0) |
| Self-reported eyesight, n (%)  Excellent  Good  Fair  Poor  Very poor/ partially sighted/ blind | 21 (10.4)  124 (61.4)  46 (22.8)  8 (4.0)  3 (1.5) | 3 (13.6)  12 (54.5)  7 (13.6)  0  0 |
| Fall (last 12 months), n (%)  Never  Once  Twice or more | 118 (58.4)  38 (18.8)  46 (22.8) | 13 (59.1)  5 (22.7)  4 (18.2) |
| EQ-5D, mean (SD) | 0.77 (0.25) | 0.69 (0.29) |
| EQ-~VAS, mean (SD) | 67.59 (18.98) | 69.32 (15.61) |
| Risk of depression, n (%)  Low risk (GDS<2)  At risk | 121 (59.9)  81 (40.1) | 14 (63.6)  8 (36.3) |
| PASE, mean (SD) | 71.02 (44.63) | 65.28 (38.57) |
| Short FES-I, mean (SD) | 14.86 (6.01) | 14.82 (4.95) |
| FRAT  Lower risk of falling (0-2)  Greater risk of falling (≥3) | 115 (56.9)  87 (43.1) | 12 (54.5)  10 (45.5) |
| BBS, mean (SD) | 42.68 (10.21) | 42.91 (8.77) |

*Frequencies are greater than sample size given that some participants owned multiple types of technology. EQ-5D-5L- EuroQol 5-Dimension 5-level; EQ-VAS- EuroQol visual analogue scale; GDS- Geriatric depression score; Short FES-I- International short falls efficacy scale; FRAT- Falls risk assessment tool; BBS- Berg balance scale

**Supplementary Table 3:** Follow-up outcome data stratified by treatment group

| **Outcome** | **Timepoint** | **Intervention (n=91)** | **Control (**6 weeks: n=87; 12 weeks n=89) | **Total (**6 weeks: n=178; 12-weeks n=180) |
| --- | --- | --- | --- | --- |
| **BBS** | *6-weeks* | 48.26 (8.18) | 43.91 (10.49) | 46.13 (9.60) |
|  | *12-weeks* | 50.27 (7.08) | 43.38 (10.97) | 46.92 (9.79) |
| **EQ-5D-5L** | *6-weeks* | 0.83 (0.21) | 0.70 (0.31) | 0.77 (0.27) |
|  | *12-weeks* | 0.87 (0.18) | 0.69 (0.33) | 0.78 (0.28) |
| **EQ-VAS** | *6-weeks* | 73.34 (19.09) | 67.66 (19.16) | 70.56 (19.28) |
|  | *12-weeks* | 73.21 (19.97) | 66.65 (18.91) | 69.97 (19.68) |
| **PASE** | *6-weeks* | 86.75 (57.54) | 71.98 (46.26) | 79.53 (52.70) |
|  | *12-weeks* | 83.54 (52.97) | 73.32 (63.84) | 78.49 (58.66) |
| **Short FES-I** | *6-weeks* | 12.09 (4.51) | 14.86 (6.00) | 13.44 (5.46) |
|  | *12-weeks* | 10.67 (4.11) | 14.57 (6.31) | 12.60 (5.65) |
| **5-STS (s)** | *6-weeks* | 26.68 (17.10) | 23.55 (15.06) | 25.63 (16.58) |
|  | *12-weeks* | 20.33 (12.63) | 30.62 (19.61) | 25.36 (17.16) |
| **Pain scale** | *6-weeks* | 23.57 (26.70) | 32.83 (29.14) | 28.10 (28.22) |
|  | *12-weeks* | 20.60 (24.40) | 29.96 (30.03) | 25.23 (27.65) |
| **Fatigue scale** | *6-weeks* | 33.13 (29.37) | 36.98 (28.77) | 35.00 (29.06) |
|  | *12-weeks* | 31.19 (26.31) | 37.08 (29.54) | 34.10 (28.03) |

**Supplementary Table 4:** Breakdown of EQ-5D-5L over the 12-week intervention

|  | **Baseline n (%)** | | **Week 12 n (%)** | |
| --- | --- | --- | --- | --- |
| **Dimension** | **KOKU** | **Control** | **KOKU** | **Control** |
| **Mobility** | | | | |
| No problems | 37 (38.1) | 25 (27.2) | 53 (57.6) | 29 (32.6) |
| Slight problems | 27 (27.8) | 12 (13.0) | 22 (23.9) | 15 (16.9) |
| Moderate problems | 25 (25.8) | 35 (38.0) | 15 (16.3) | 32 (36.0) |
| Severe problems | 8 (8.2) | 20 (21.7) | 1 (1.1) | 12 (13.5) |
| Unable to walk about | - | - | - | 1 (1.1) |
| **Self-care** | | | | |
| No problems | 65 (67.0) | 59 (64.1) | 71 (77.2) | 53 (59.6) |
| Slight problems | 15 (15.5) | 13 (14.1) | 11 (12.0) | 17 (19.1) |
| Moderate problems | 16 (16.5) | 16 (17.4) | 8 (8.7) | 12 (13.5) |
| Severe problems | 1 (1.0) | 4 (4.3) | - | 6 (6.7) |
| Unable to wash or dress | - | - | 1 (1.1) | 1 (1.1) |
| **Usual activities** | | | | |
| No problems | 50 (51.5) | 40 (43.5) | 60 (65.2) | 38 (42.7) |
| Slight problems | 23 (23.7) | 13 (14.1) | 19 (20.7) | 17 (19.1) |
| Moderate problems | 16 (16.5) | 22 (23.9) | 11 (12.0) | 25 (28.1) |
| Severe problems | 8 (8.2) | 16 (17.4) | 1 (1.1) | 8 (9.0) |
| Unable to do usual activities | - | 1 (1.1) | - | 1 (1.1) |
| **Pain/ discomfort** | | | | |
| No pain/ discomfort | 31 (32.0) | 23 (25.0) | 47 (51.1) | 28 (31.5) |
| Slight pain/ discomfort | 26 (26.8) | 24 (26.1) | 19 (20.7) | 20 (22.5) |
| Moderate pain/ discomfort | 31 (32.0) | 30 (32.6) | 22 (23.9) | 21 (23.6) |
| Severe pain/ discomfort | 6 (6.2) | 11 (12.0) | 2 (2.2) | 19 (21.3) |
| Extreme pain/ discomfort | 3 (3.1) | 4 (4.3) | 1 (1.1) | 1 (1.1) |
| **Anxiety/ depression** | | | | |
| Not anxious/ depressed | 51 (52.6) | 45 (48.9) | 58 (63.0) | 40 (44.9) |
| Slightly anxious/ depressed | 20 (20.6) | 15 (16.3) | 18 (19.6) | 25 (28.1) |
| Moderately anxious/ depressed | 20 (20.6) | 29 (31.5) | 13 (14.1) | 17 (19.1) |
| Severely anxious/ depressed | 5 (5.2) | 2 (2.2) | 1 (1.1) | 7 (7.9) |
| Extremely anxious/ depressed | 1 (1.0) | 1 (1.1) | 1 (1.1) | - |

**Supplementary Table 5:** Theoretical framework for acceptability (TFA) based on use of KOKU

| **TFA component** | **Mean (SD) score** |
| --- | --- |
| Affective attitude | 4.44 (0.56) |
| Burden | 3.22 (1.50) |
| Ethicality | 3.85 (0.79) |
| Perceived effectiveness | 4.43 (0.69) |
| Intervention coherence | 4.33 (0.82) |
| Self-efficacy | 4.49 (0.72) |
| Opportunity costs | 3.50 (1.42) |
| General acceptability | 4.64 (0.48) |

**Supplementary Table 6:** Unit costs of primary, and secondary healthcare resources (2023/24 £).

| Service type | Unit costs | Source |
| --- | --- | --- |
| GP surgery | £45 | PSSRU 2024 (general practitioner cost per surgery consultation lasting 10 minutes, including overhead and direct health care staff costs, including training). |
| Outpatient visits | £250 | National Cost Collection 2023/24 (National average unit cost of outpatient attendances). |
| Day cases | £1,031 | National Cost Collection 2023/24(National average unit cost of day cases). |
| Accident & emergency attendances | £273 | National Cost Collection 2023/24 (National average unit cost of A&E attendances not leading to admission). |
| Inpatients admissions (Long stay) | £5,134 | National Cost Collection 2023/24 (National average unit cost of elective inpatients, long stay (FCEs)). |
| Inpatients admissions (Short stay) | £792 | National Cost Collection 2023/24 (National average unit cost of elective inpatients, short stay (FCEs)). |

Note: PSSRU Personal Social Services Research Unit.

**Supplementary Table 7:** Sensitivity Analysis

| **Sensitivity analysis** | **Incremental cost, £ (SE)** | **95% CI** | **Incremental QALYs (SE)** | **95% CI** | **ICER** |
| --- | --- | --- | --- | --- | --- |
| (1) Complete case analysis | -87.33 (79.22) | [-268.79, 32.07] | 0.019 (0.007) | [0.005, 0.033] | Dominant |
| (2) Intervention cost without tablet | -78.99 (68.18) | [-234.54, 24.22] | 0.021 (0.007) | [0.007, 0.034] | Dominant |
| (3) 10% lower health & social care costs | -52.38 (61.37) | [-192.38, 40.50] | 0.020 (0.007) | [0.004, 0.033] | Dominant |
| (4) 10% higher health & social care costs | -73.58 (75.00) | [-244.69, 39.94] | 0.020 (0.008) | [0.003, 0.035] | Dominant |
| (5) Tablet assumption: 1 in 4 patients (£20/patient) | -58.98 (68.19) | [-214.54, 44.22] | 0.020 (0.007) | [0.004, 0.033]] | Dominant |

***Note:*** *Estimates are based on non-parametric bootstrapping (10,005 replications). For multiply imputed analyses, 15 imputed datasets were used with 667 bootstrap replications per dataset. Costs are reported in GBP (£) using the most recent NHS unit cost sources. Dominant indicates lower costs and higher QALYs*

**Supplementary Table 8a:** Participant characteristics (older adults who received KOKU intervention) who completed an interview

| **Participant** | **Age group** | **Gender** | **Ethnicity** | **Geographic location** |
| --- | --- | --- | --- | --- |
| 01 | 60-64 | Female | Black/ Black British (Caribbean) | Manchester Central |
| 02 | 65-69 | Female | White British | Manchester Central |
| 03 | 85-89 | Female | Black/ Black British (Caribbean) | Manchester Central |
| 04 | 70-74 | Female | White British | Bury |
| 05 | 60-64 | Male | Hong Kong | Bury |
| 06 | 60-64 | Female | White British | Manchester Central |
| 07 | 80-84 | Female | White British | Stockport |
| 08 | 80-84 | Female | White British | Manchester Central |
| 09 | 90-94 | Female | White British | Stockport |
| 10 | 85-89 | Female | White British | Bury |
| 11 | 70-74 | Male | White British | Salford |
| 12 | 75-79 | Female | White British | Bury |
| 13 | 65-69 | Male | White British | Stockport |

**Supplementary Table 8b:** Participant characteristics (Social care staff) who completed an interview

| **Participant** | **Role** | **Provider Type** |
| --- | --- | --- |
| 01 | Operations Manager | Social Care Charity |
| 02 | Carers Service Project Worker | Social Care Charity |
| 03 | Dementia Advisor | Social Care Charity |
| 04 | Ageing in Place Development Worker | Not-for-profit company |
| 05 | Team Lead | Council |
| 06 | Project Support Officer | Council |
| 07 | Frailty Coach | Health and Social Care |
| 08 | Senior Manager | Social Care Charity |
| 09 | Manager of Volunteer Services | Social Care Charity |
| 10 | Age Friendly Project Manager | Community-focused Housing Association |

**Supplementary Table 9:** Example quotes to reflect themes from reflexive thematic analysis of interviews

| Theme | Quotes |
| --- | --- |
| *High ease of use increases confidence, motivation and engagement* | “Easy, very easy. I found it easy and I think it was incredible as well. It's good. It was very good” (Participant 4)  “I didn't have any issues with the exercises, as they were very easy and simple. I’ve done some more difficult exercises in the past, and I think the KOKU exercises are suitable for older people or those who are recovering from illness” (Participant 5)  “Well, the exercises, they were very well, you know, sort of explained and obviously [the e-coach] showed you how you know what to do to, you know, to obviously to make it, you know, make it safe” (Participant 6)  “As time goes on. I mean, it's all right. I can do those exercises. Yeah. Yeah, I've done 15 of these. Yeah, right on to the next one. Because as I say, as time goes on, they do become a bit easier. They do become normalised” (Participant 1)  “I think the program is very easy to control. I can do it whenever I want – whether it's Monday morning, afternoon, or evening. It only takes a few minutes to complete the exercises, and if I have more time that day, I can repeat them. There's no time limit, so I can do it whenever suits me. It’s very flexible and easily accessible. Even if it's raining outside, I can still exercise indoors” (Participant 5)  “It didn’t say how many repetitions to do. I've tried to download it again this morning to refresh my memory and I can't. I well won't come up on the App Store, so I haven't been able to refresh my memory. I've got my notes that I made, but just thinking about it, it didn't, it wasn't specific enough for me in the fact of how many repetitions or how long we had to do them and so it wasn't, it wasn't specific enough in a way… Well, I think in the past when I've done exercises it's been more how many repetitions and how many sort of counts to do an exercise too. So I suppose it would be both really” (Participant 7) |
| *Improving health, wellbeing and functioning through routine KOKU use* | “I feel that the difference is in my muscles. They’ve become stronger, which makes it easier for me to move around. The more I do the exercises, the stronger my muscles get. Yes, my leg muscles are stronger. It’s helped with my balance as well” (Participants 5)  “I decided to do it because I’ve got osteoporosis and I thought it might help me … I feel more comfortable moving around. I find it easier to go upstairs now, and when I get in the bath, I feel more steady… It definitely helped improve my movement – though I feel a bit stiff now because I haven’t been doing much recently.” (Participant 4)  “It has given me more confidence…I’ve now started going, I do line dancing… somebody actually commented last week that I’m getting up more, I’m trying, you know, trying them all. (Participant 6).  “Once you do the exercises, your mood changes, you know, you feel happier in yourself” (Participant 3).  “Well, it stopped me from reaching for something I shouldn't be eating. Because it would say, oh, this is good for you. Yes, drink more water. And would you believe it picked up a glass of water and I drank it” (Participant 1) |
| *Tailoring challenge to ability* | “Well, I found it. It’s quite easy because I go to a Pilates class also… I didn’t find them difficult [even as time progressed]… there may be some sort of more challenging exercises that would be helpful” (Participant 10)  “….to be personal for everyone else, I think having more exercises on instead of the one, two, three, that's all there was. Maybe if they did four, five, six on the first block, then increase the number gradually…” (Participant 4)  “I think again [the games] could do with a bit more of a progression on it as well. Bit more harder. Or would you like to try something slightly different or with a bit more content in it, instead of five hazards…we've got 10 hazards here now. Which ones can you see? You know?” (Participant 1) |
| *Simple, safe and beneficial* | “I think its just like the relevance of KOKU for our service users and for the carers as well who are supporting people who have had falls or themselves are getting older and need to sort of use it at the earliest stage rather than when its too late, so yeah, its just really relevant for all our service users” (Staff, 02)  “Its been really good have all those options, because everybody learns differently, don’t they? Everyone, some people would just focus on the character, how they’re moving, and some people will read. So having all those options, it kind of caters for everybody really. So depending on how you learn best, you can just follow the prompts” (Staff 07)  “I think the KOKU app is a very good tool not just for myself, maybe not just for our team, even for our wellbeing staff.. I think it has proven that the app is really easy to use… because all the explanation and guidance on the app is really clear, so both of their safety can be protected because there are different safety advice before start doing the exercise. So it is really safe for people to use the app even” (Staff 03) |
| *Supporting falls prevention in a digital age* | “So anything that can prevent [falls and hospital admissions] from happening and making us stronger and healthier for longer, I'm one hundred percent for the idea that a free tablet really, really went down with our participants, our cohort because of course being in Gorton, being impoverished, I don't know if that's the correct term, but you know, not all white, affluent that could afford their own tech equipment…” (Staff 04)  “Sometime people do have a paper leaflet type of thing, but I don’t know. People aren’t always good at using those, as you could be just putting that in a drawer. With KOKU, its interactive, its there right in front of you as its something visual. You get into a habit of doing it. I think it’s a good habit thing, isn’t it?” (Staff 08)  “First impressions that I had when I was first showing Koku would be that this would be brilliant for all the patients that are housebound that are unable to access community services and care homes…so all those patients that are in a care home that don't like to socialize and do the group classes with them, they can be done on a one to one basis with carers and family” (Staff 07)  “It's rewarding like you get something, right, nutrition, and then you get ‘ding!’ your little bell, which is which is always good to have that positive reinforcement no matter what you are doing” (Staff 01)  “It feels like you have somebody and you're interacting there. It’s that interaction even just on that very base level where it's direct and there's a response” (Staff 09)  “I was just going to say in terms of accessibility, the biggest limitation that I see is that it's only available on one device, which is a tablet and everyone has mobile phones. People have laptops, but if they don't have a tablet and they're going to struggle to be able to use it” (Staff 01)  “For the character I do not have much idea how to like improve it but what I’m concerned (about) is also diverse older people. Some are Black, like Asian, some are White British, so we need to think about different ethnic multi groups using this health app” (Staff 10)  “I was just going to say in terms of accessibility, the biggest limitation that I see if that it’s only available on one device, which is a tablet and everyone has mobile phones. People have laptops, but if they don’t have a tablet and they’re going to struggle to be able to use it” (Staff 01) |
| *Empowering self-management to reduce NHS demand* | “If we could introduce this in the home environment in a time where their pain is optimized and they’re in less pain at times and able to exercise then potentially we could try and benefit their wellbeing by doing KOKU and then improve this would improve their mood, improve the general health, then make better lifestyle choices. And then take pressure off the NHS as well so its also more cost-effective” (Staff 07)  “Our role is a prevention intervention service to try and keep people out of services in the NHS, so we will go in and try and support people to take responsibility for their own health and well-being, and this is an amazing app to be able to get people to do that” (Staff 05)  “There's an awful lot of benefits to it that might not be so obvious and we see we see that there is a knock on effect with very, very small changes. There's a knock on effect to mood… We know the benefits of something like [this]… Obviously on mobility and joints… and the psychological well-being benefits, mood lifters” (Staff 09)  “Two people [we had] where they'd not had the confidence to go to a group and want to go to an exercise group…that was too daunting for them. However, once they'd started to do the exercises at home, they enjoyed it so much, felt the benefit of it, and then went on to a group and then got another friend involved in going to a group. So it gave them confidence to do those extra [things]” (Staff 06)  “I think just gave them a tiny bit more confidence to maybe join that walk because we have a sort of a beginners walking group, which is a good lead on from the koku and perhaps the social aspect of being able to discuss it together. Yeah. And being in the team and even though they didn't actually say that it's more of an observation seeing them… they're confident [more] confident now” (Staff 04)  “I think getting people together for our lot, I think that would work for them because they would come for the natter and the exercise and it would get them out of the house and they would see each other… I think that would encourage them and again hot the socialisation sort of loneliness problem on the head, checking up on them because nobody does anything unless they’re checked up on really do they? We’re all guilty of that” (Staff 04) |

**Supplementary Figure 1:** Short version of user experience questionnaire results for 90 participants using KOKU compared to benchmark data from 21175 individuals

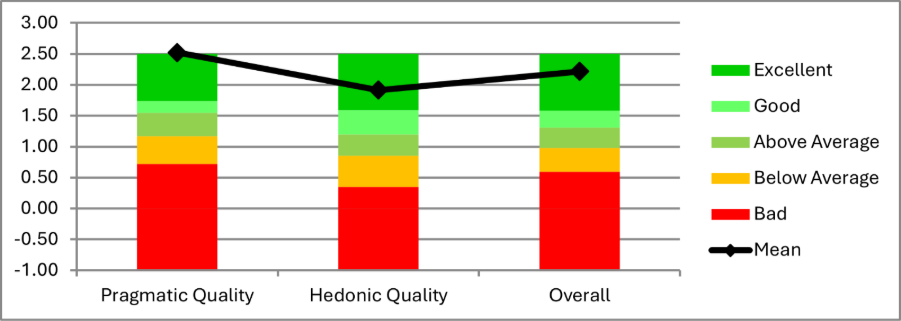

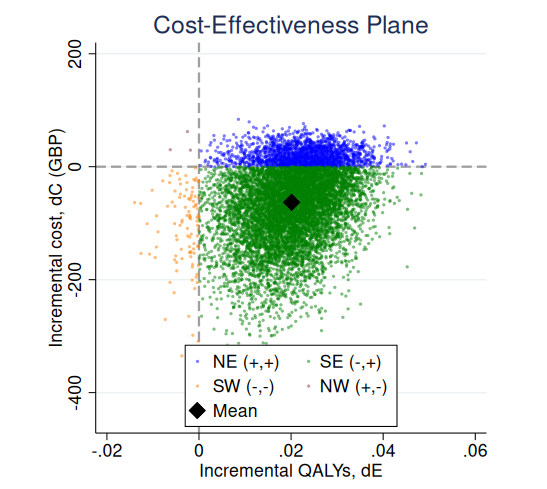
**Supplementary Figure 2:** Cost-effectiveness plane for KOKU vs control group

***Note:*** *The cost-effectiveness plane shows bootstrapped incremental costs (dC) and QALYs (dE) for the intervention versus usual care. Each point represents one bootstrap replication. Blue, green, orange, and red points represent the north-east, south-east, south-west, and north-west quadrants, respectively. The black diamond indicates the mean incremental cost and effect.*

**Supplementary Figure 3:** Incremental Net Monetary Benefit at £20,000 and £30,000 Willingness-to-Pay Thresholds

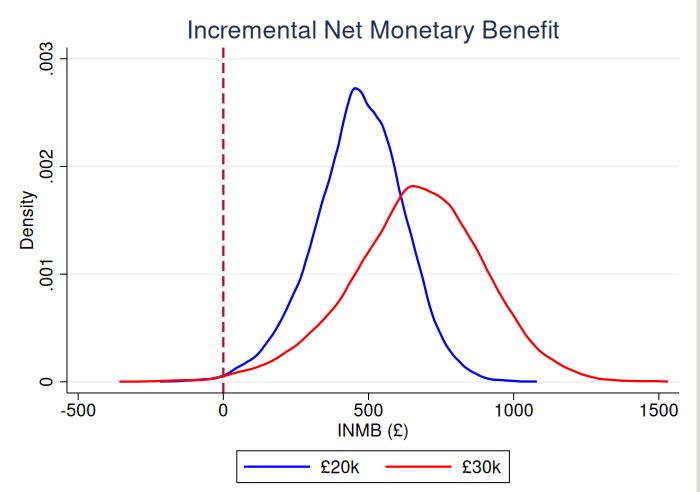

***Note:*** *The figure shows the bootstrap distribution of incremental net monetary benefit (INMB) for the intervention compared with usual care at willingness-to-pay thresholds of £20,000 and £30,000 per QALY gained. Values to the right of zero indicate the intervention is cost-effective*

**Supplementary Figure 4:** Cost-effectiveness acceptability curve (Outcome: Quality adjusted life years (QALY))

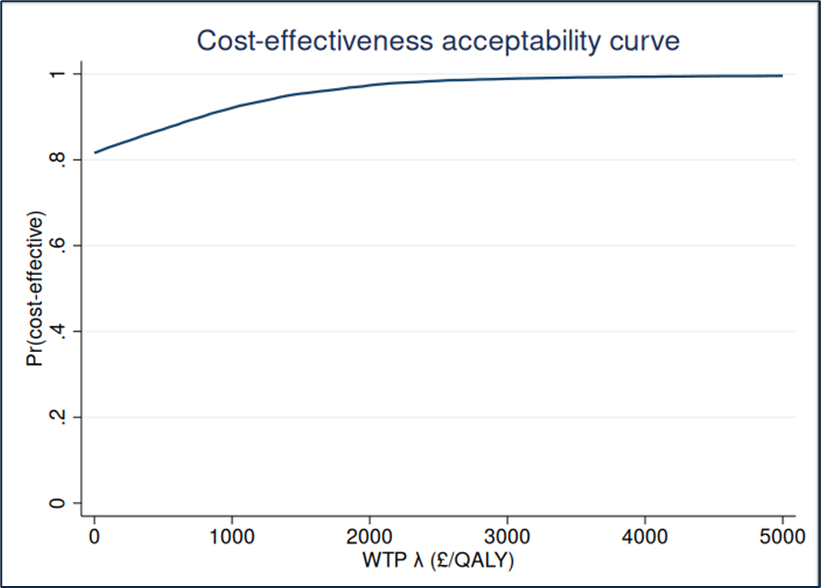

***Note****: The cost-effectiveness acceptability curve shows the probability that the intervention is cost-effective compared with usual care across a range of willingness-to-pay (WTP) thresholds per quality-adjusted life year (QALY). The curve is based on bootstrapped estimates (10,005) of incremental costs and QALYs*
